# Association Between Person-Centered Care Quality and Advance Care Planning Participation in Patients Undergoing Hemodialysis: A Multicenter Cross-Sectional Study

**DOI:** 10.1101/2023.12.15.23300045

**Authors:** Yusuke Kanakubo, Noriaki Kurita, Mamiko Ukai, Tetsuro Aita, Ryohei Inanaga, Atsuro Kawaji, Takumi Toishi, Masatoshi Matsunami, Yu Munakata, Tomo Suzuki, Tadao Okada

## Abstract

**Rationale & Objective:** Person-centered care (PCC), which incorporates patients’ preferences and values not only for medical care but also for their life, in decision making has been proposed for promoting advance care planning (ACP) among patients with kidney failure.

However, how variations in PCC affect ACP participation remain unclear. Therefore, we examined variations in PCC across facilities and examined the association between PCC and ACP participation.

**Study Design:** Multicenter cross-sectional study.

**Setting & Participants:** Japanese adults receiving outpatient hemodialysis at six dialysis centers.

**Exposures:** PCC was measured using the 13-item Japanese version of the Primary Care Assessment Tool-short form.

**Outcome:** ACP participation as defined by discussion with the attending physician or written documentation or notes regarding treatment preferences.

**Analytical Approach:** A general linear model was used to examine the correlates of the quality of PCC. Modified Poisson regression models were used to examine the associations of ACP participation.

**Results:** A total of 453 individuals were analyzed; 26.3% participated in ACP. Compared to respondents with no usual source of care (USC), higher PCC was associated with greater ACP participation in a dose-response manner (vs. no USC, adjusted prevalence ratios for the first to fourth quartiles: 1.36, 2.31, 2.64, and 3.10, respectively). Among the PCC sub-domains, first contact, longitudinality, comprehensiveness (services provided), and community orientation were particularly associated with ACP participation. There was a maximum of 12.0 points of facility variation in the quality of PCC.

**Limitations:** Possible reverse causation and unmeasured confounders.

**Conclusions:** High PCC quality was associated with ACP participation. The substantial disparity in PCC between facilities provides an opportunity to revisit the quality improvement in PCC.

## Introduction

Advance care planning (ACP) is a process involving discussion and documentation of the patient’s wishes regarding goals and preferences for future medical treatment and care, especially for patients who may lose physical capacities and communication skills in the future. ^1,2^ The importance of ACP in end-of-life care is highlighted by benefits such as fewer invasive medical procedures, improved quality of life, and fewer hospitalizations. ^1,3–5^ The relevance of ACP for patients on dialysis is particularly high because of the unique decision-making requirements, such as dialysis withdrawal, at the end of life. According to a United States Renal Data System report, dialysis withdrawal, including that secondary to acute conditions, accounts for 17% of all deaths. ^6^ Although dialysis providers play an important role in helping patients reconstruct what they imagine and desire for their future treatment as their disease progresses, ^7^ there is insufficient evidence on how dialysis providers should implement ACP.

Previous studies suggested that facilitators of ACP implementation should include staff training and clarifying core values of one’s life for patients on dialysis through dialogue. ^8–10^ Such “person-centered care” that focuses on patient preferences, needs, and values not only related to healthcare but also to one’s whole life is emphasized in the field of nephrology. ^11,12^ Patients undergoing hemodialysis are likely to rely on their dialysis provider for person-centered care about day-to-day problems because they visit the same dialysis provider thrice a week. ^13^ Thus, for these patients, the dialysis provider can be viewed as a usual source of care (USC) providing primary care. ^14^ However, whether person-centered care for patients on dialysis is indeed associated with ACP preparedness and participation has not been adequately examined. Focusing on person-centered care quality will allow us to examine the association between such care and ACP preparedness as demonstrated in outpatients and patients with chronic disease on homecare. ^15,16^ Additionally, it is possible to identify and visualize variations in person-centered care across facilities and improve it, considering the reality that some dialysis providers disagree with making time or going beyond their regular job description. ^11,17^

Therefore, we conducted a multicenter cross-sectional study to clarify whether person-centered care is associated with ACP preparedness among Japanese patients on hemodialysis. We also assessed variations in person-centered care across dialysis facilities.

## Methods

### Study design and subjects

This was a multicenter study conducted across six medical institutions in Chiba, Tokyo, and Kanagawa. The study enrolled adult patients undergoing maintenance hemodialysis at the participating facilities. Patients who medical professionals thought were unable to respond on their own to this survey were excluded. Before participation, the research participants were provided with information about the study and asked to express their consent to participate. Written informed consent was obtained from all participants. We requested the participants who provided informed consent to respond to a questionnaire. The questionnaires were collected by staff, ensuring anonymity in content discernment. Additionally, the attending physicians extracted data related to dialysis from the medical records. As an incentive, respondents were offered monetary gratification in the form of a 500-yen (equivalent to US$ 4.1, based on the exchange rate of 122 yen to US$ 1 at that time) gift card. Data were collected from April 2022 to February 2023. This study adhered to the principles enshrined in the Declaration of Helsinki and received approval from the Institutional Review Board at Fukushima Medical University (Approval No. number ippan2021-292).

### Outcome: Participation in ACP

Aligned with the administration of a treatment preference questionnaire among patients with kidney failure in the United States, ^18^ the subsequent instructional statement was provided as follows: “This section asks about thoughts on your health care if you were to become very sick in the future.” Subsequently, the ensuing query was presented as: “Have you thought about the kinds of treatments you would want or not want if you were to become very sick and were unable to speak in the future? (check all items that apply).” Multiple options were offered, encompassing the ensuing responses: “I have not thought about this,” “I have thought about this, but have not talked about it to a family member, others (including a friend), or my doctor,” “I have talked about this with a family member or others (including a friend),” “I have talked about this with my doctor,” or “I have written a document or memo about my preferences.” The inclusion criterion for ACP participation was operationalized when participants selected either the response option “I have talked about this with my doctor” or “I have written a document or memo about my preferences” because recent studies concerning ACP interventions have employed a combination of effective communication and the formalization of advance directives. ^15,16,19,20^

### Exposure: Patient experience of person-centered care

To assess the quality of person-centered care, we used the 13-item Japanese version of the Primary Care Assessment Tool-short form (JPCAT-SF). ^21^ The full version of the JPCAT-SF is a modified version of the Primary Care Assessment Tool, which was developed by B. Starfield, L. Shi, and colleagues at the Johns Hopkins Primary Care Policy Center and comprehensively assesses the primary care characteristics, to fit the Japanese context. ^22,23^

It is one of the quality indicators of healthcare measuring person-centered care, which is the cornerstone of primary care, by asking about events that patients experience during the care process. ^23,24^ The JPCAT-SF consists of the same six domains as the JPCAT, which represent five primary care attributes: first contact (2 items), longitudinality (2 items), coordination (3 items), comprehensiveness (2 items for services available and 2 items for services provided), and community orientation (2 items). The JPCAT-SF has good internal consistency reliability (Cronbach’s α = 0.77 for total scores and Cronbach’s α > 0.76 for each domain score) and excellent criterion validity (Pearson correlation coefficient with the original 29-item JPCAT and the overall rating for usual care facilities: 0.94 and 0.43, respectively). ^21^ Details of the JPCAT-SF items and domains are shown in Supplementary Items S1 and S2.

Subjects chose “Yes” or “No” to the first question of the JPCAT-SF “Is there a doctor that you usually go to if you are sick or need advice about your health?” If they chose “Yes,” they were considered as having a USC. Next, scores on the JPCAT-SF items were calculated for those having a USC. The subjects rated each item on a 5-point Likert scale. Scores for each domain were calculated from 0 to 100 according to the established algorithm. The JPCAT-SF total score was calculated as the mean score for each domain. Higher scores indicated higher quality of person-centered care.

### Data collection and covariates

Age, gender, education, household income, questions regarding ACP, and the JPCAT-SF were collected by a self-administered questionnaire.

Data on the underlying kidney disease, type of hemodialysis therapy (hemodialysis or hemodiafiltration), comorbidities, and dialysis duration were collected by attending physicians at each dialysis facility from the patient’s medical records. Comorbidities were scored based on the modified Charlson comorbidity index (CCI) for patients on dialysis. ^25^ Age, gender, education, household income, modified CCI, dialysis duration, and dialysis facility were chosen as covariates.

### Statistical analysis

Statistical analyses were conducted using Stata/SE, version 17 (StataCorp, College Station, TX, USA). Patient characteristics are presented as means and standard deviations (SDs) for continuous variables and as numbers and proportions for categorical variables.

To estimate the association between the JPCAT-SF score and ACP participation, a modified Poisson regression model was fitted. ACP participation was considered the outcome variable, while the JPCAT-SF total score, age, sex, education, household income, modified CCI, dialysis vintage, and dialysis facility were included as explanatory variables. The choice of the model aimed to directly estimate the prevalence ratio (PR) of the primary outcome variable with a non-rare proportion. ^26^ The JPCAT-SF total score was classified into five categories. The reference category comprised participants who reported not having a USC. ^27^ The remaining four categories were determined based on the JPCAT-SF total score, aiming to achieve equal participant numbers in each group.

Similar to the main analysis, modified Poisson regression models adjusted for the aforementioned covariates were used to estimate the associations between the five categorized JPCAT-SF subdomains (with no USC as the reference) and ACP participation. As supplementary analyses, the associations between ACP participation and both the JPCAT-SF total and subdomain scores as continuous values were evaluated. This was done using the aforementioned model and covariates and limited to patients who reported having a USC. Furthermore, a general linear model was applied to patients who reported having a USC to assess differences in JPCAT-SF total scores across different dialysis facilities, with adjustment for the aforementioned covariates. Finally, the model was used to estimate the predicted mean JPCAT-SF total score for the entire analysis population using Stata’s *margin* command, ^28^ assuming that each individual belonged to a specific facility.

To account for covariates with missing data, we employed the multiple imputation with chained equations method, generating 20 imputations to impute the missing data. This approach assumed that the missing data occurred at random. For each analysis, we used a two-tailed significance level of P < 0.05.

## Results

### Study flow and participant characteristics

Of the 651 patients undergoing hemodialysis at the six facilities, 484 (74.3%) participated in this study. Of these, 25 and 6 patients were excluded because of missing data on ACP participation and JPCAT total score, respectively, resulting in a total of 453 patients for the analysis (Fig. 1).

**Figure 1.**
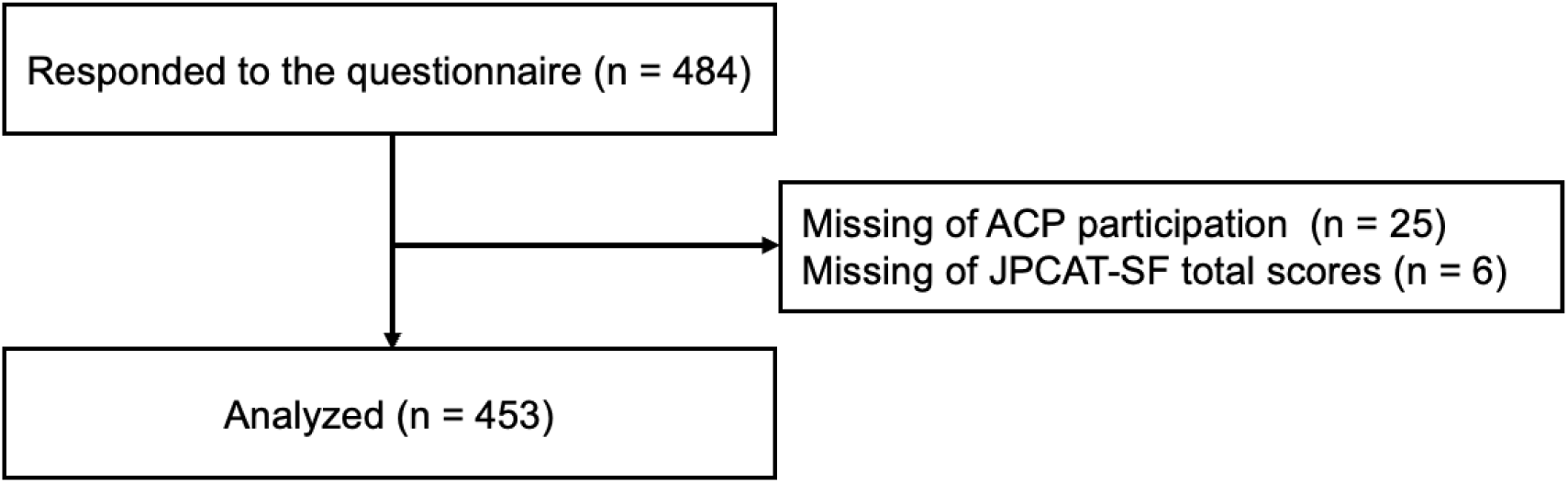
Patient flowchart.

**Fig. 2.**
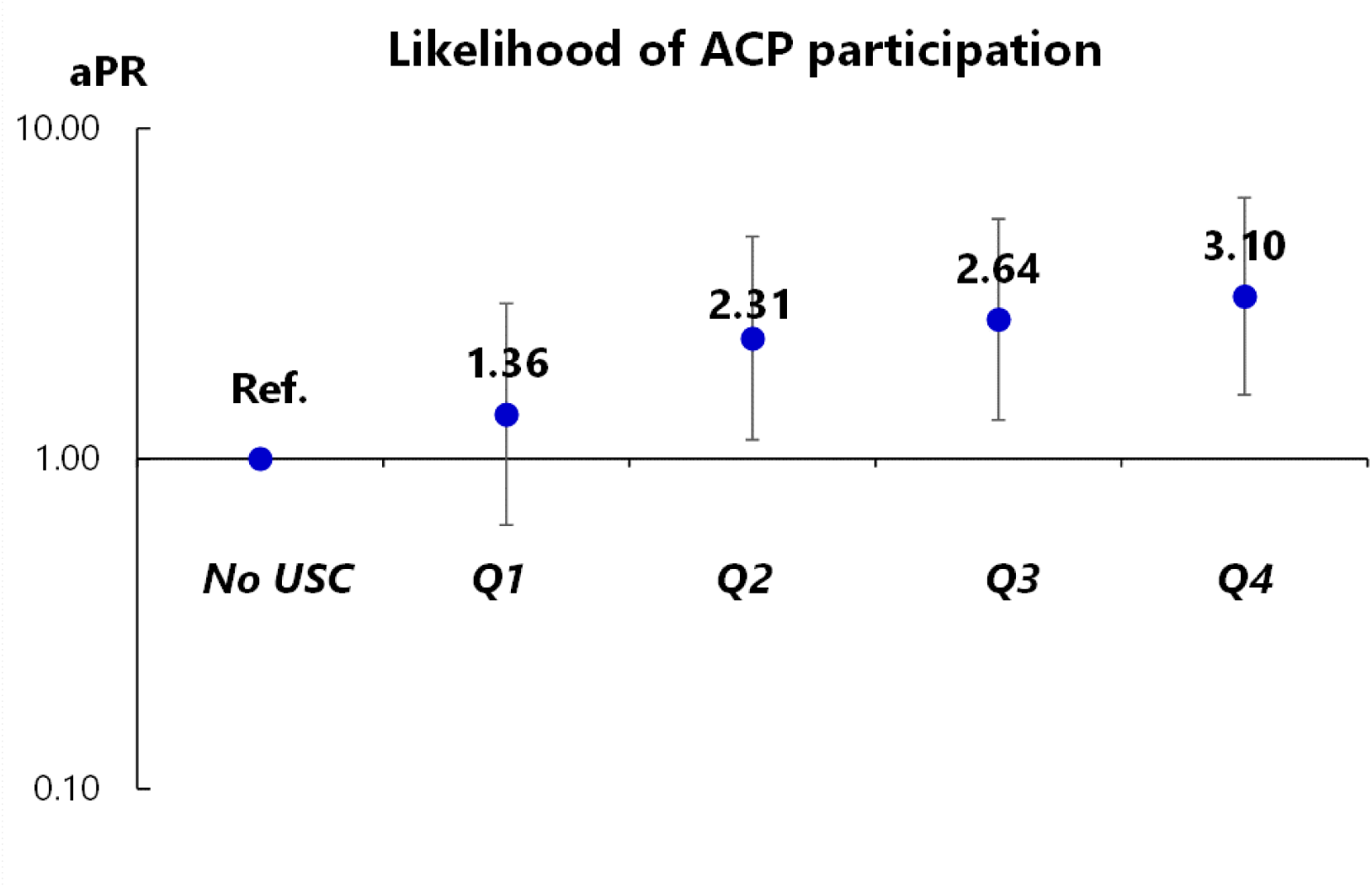
Association between person-centered care, as measured by the JPCAT-SF total score, and ACP participation. USC = usual source of care Notes: Adjusted for age, sex, education, household income, comorbidities, dialysis vintage, and dialysis facility. Error bars indicate 95% confidence intervals. JPCAT-SF total score quartiles: Q1, 0.0–50.0; Q2, 50.0–60.0; Q3, 60.0–70.0; Q4, 70.0–100.0.

Table 1 shows the characteristics of the study population.

**Table 1.**
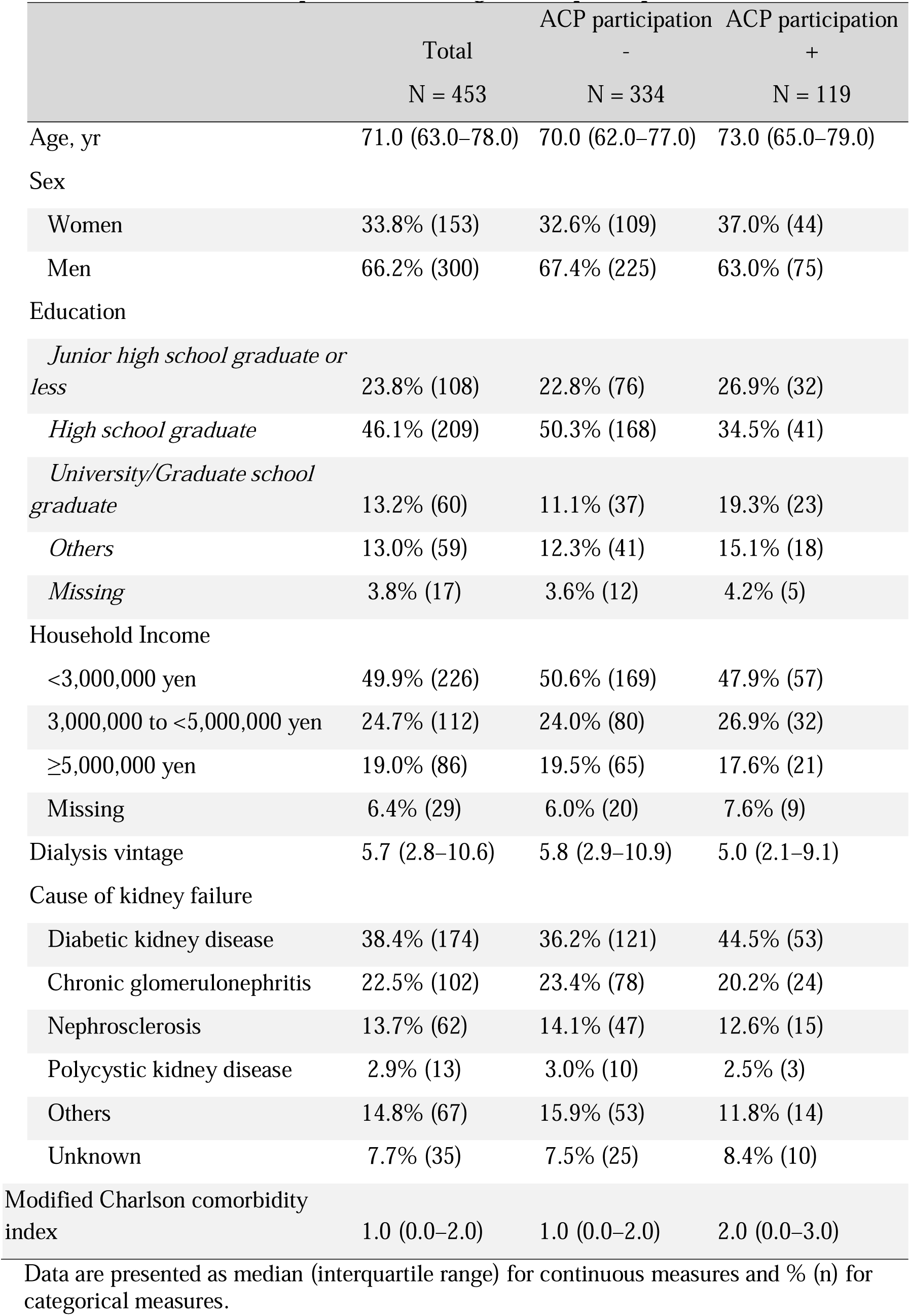
Characteristics of patients according to ACP participation (n = 453)

|  | Total<br>N = 453 | ACP participation<br>-<br>N = 334 | ACP participation<br>+<br>N = 119 |
| --- | --- | --- | --- |
| Age, yr | 71.0 (63.0–78.0) | 70.0 (62.0–77.0) | 73.0 (65.0–79.0) |
| Sex |  |  |  |
| Women | 33.8% (153) | 32.6% (109) | 37.0% (44) |
| Men | 66.2% (300) | 67.4% (225) | 63.0% (75) |
| Education |  |  |  |
| *Junior high school graduate or less* | 23.8% (108) | 22.8% (76) | 26.9% (32) |
| *High school graduate* | 46.1% (209) | 50.3% (168) | 34.5% (41) |
| *University/Graduate school graduate* | 13.2% (60) | 11.1% (37) | 19.3% (23) |
| *Others* | 13.0% (59) | 12.3% (41) | 15.1% (18) |
| *Missing* | 3.8% (17) | 3.6% (12) | 4.2% (5) |
| Household Income |  |  |  |
| <3,000,000 yen | 49.9% (226) | 50.6% (169) | 47.9% (57) |
| 3,000,000 to <5,000,000 yen | 24.7% (112) | 24.0% (80) | 26.9% (32) |
| ≥5,000,000 yen | 19.0% (86) | 19.5% (65) | 17.6% (21) |
| Missing | 6.4% (29) | 6.0% (20) | 7.6% (9) |
| Dialysis vintage | 5.7 (2.8–10.6) | 5.8 (2.9–10.9) | 5.0 (2.1–9.1) |
| Cause of kidney failure |  |  |  |
| Diabetic kidney disease | 38.4% (174) | 36.2% (121) | 44.5% (53) |
| Chronic glomerulonephritis | 22.5% (102) | 23.4% (78) | 20.2% (24) |
| Nephrosclerosis | 13.7% (62) | 14.1% (47) | 12.6% (15) |
| Polycystic kidney disease | 2.9% (13) | 3.0% (10) | 2.5% (3) |
| Others | 14.8% (67) | 15.9% (53) | 11.8% (14) |
| Unknown | 7.7% (35) | 7.5% (25) | 8.4% (10) |
| Modified Charlson comorbidity index | 1.0 (0.0–2.0) | 1.0 (0.0–2.0) | 2.0 (0.0–3.0) |

Data are presented as median (interquartile range) for continuous measures and % (n) for categorical measures.

The mean age was 69 years (SD, 12.3); 46.1% completed high school, and 49.9% had an annual household income of <3 million yen. The median (interquartile range [IQR]) dialysis duration was 5.7 years (2.8–10.6). Overall, 86.5% of the respondents reported having a USC.

### Association of ACP participation with person-centered care

Among those analyzed, 107 (23.6%) patients had discussed ACP with their physicians, and 34 (7.5%) had recorded it. A total of 119 (26.3%) participated in ACP.

Table 2 and Figure 2 show the association between the JPCAT-SF total score and ACP participation, demonstrating that the JPCAT-SF total score has a dose-dependent association with ACP participation: with no USC as a reference, the aPR increased from 1.36 (95% confidence interval [CI], 0.36–2.95) in the lowest quartile to 3.10 (95% CI, 1.56–6.17) in the highest quartile. A higher modified CCI was associated with higher ACP participation (per 1-point increase, aPR: 1.08 [95% CI, 1.01–1.15]).

**Table 2.** Association of ACP participation with the JPCAT-SF total score and covariates (n = 453)

|  | Adj. PR, point estimate<br>(95% CI) | P-value |
| --- | --- | --- |
| JPCAT-SF total score |  |  |
| No Usual Source of Care | Reference |  |
| Q1 | 1.36 (0.63–2.95) | 0.437 |
| Q2 | 2.31 (1.14–4.71) | 0.021 |
| Q3 | 2.64 (1.31–5.32) | 0.007 |
| Q4 | 3.10 (1.56–6.17) | 0.001 |
| Age, per 10-yr increase | 1.14 (0.996–1.31) | 0.057 |
| Women vs. Men | 0.87 (0.63–1.21) | 0.412 |
| Education |  |  |
| *Junior high school graduate or less* | Reference |  |
| *High school graduate* | 0.74 (0.49–1.1) | 0.138 |
| *University/Graduate school graduate* | 1.5 (0.93–2.43) | 0.097 |
| *Others* | 1.09 (0.67–1.78) | 0.718 |
| Household Income |  |  |
| <3,000,000 yen | Reference |  |
| 3,000,000 to <5,000,000 yen | 1.12 (0.79–1.6) | 0.523 |
| ≥5,000,000 yen | 0.99 (0.64–1.51) | 0.955 |
| Dialysis vintage, per 1-yr increase | 0.98 (0.96–1.01) | 0.143 |
| Modified Charlson comorbidity index,<br>per 1-pt increase | 1.08 (1.01–1.15) | 0.018 |

A modified Poisson regression model was fitted with adjustment for age, gender, education, household income, dialysis duration, modified Carlson Index, and the participating facilities.

Table 3 shows the association between the JPCAT-SF subdomain scores and ACP participation. Across all subdomains, categories in which patients reported receiving higher person-centered care were associated with greater ACP participation compared to those who reported not having a USC.

**Table 3.** Association of ACP participation with the JPCAT-SF subdomain scores (n = 453)

|  | Adj. PR, point estimate (95% CI) |  |  |  |  |  |
| --- | --- | --- | --- | --- | --- | --- |
|  | First Contact | Longitudinality | Coordination | Comprehensiveness (Services Available) | Comprehensiveness (Services Provided) | Community Orientation |
| JPCAT-SF scores |  |  |  |  |  |  |
| No Usual Source of Care | Reference | Reference | Reference | Reference | Reference | Reference |
| Q1 | 1.99 (0.97–4.07) | 2.54 (1.14–5.62) | 1.84 (0.91–3.73) | 2.42 (1.12–5.22) | 1.58 (0.75–3.32) | 1.81 (0.83–3.91) |
| Q2 | 1.87 (0.89–3.9) | 1.43 (0.69–2.93) | 2.91 (1.44–5.88) | 1.92 (0.92–3.99) | 2.55 (1.26–5.18) | 2.21 (1.12–4.38) |
| Q3 | 2.38 (1.18–4.78) | 2.53 (1.24–5.15) | 2.47 (1.22–5.04) | 2.53 (1.29–4.96) | 2.29 (1.13–4.66) | 2.69 (1.36–5.31) |
| Q4 | 3.56 (1.73–7.36) | 3.25 (1.64–6.42) | 2.38 (1.16–4.88) | 2.45 (1.19–5.06) | 3.03 (1.53–6.01) | 2.57 (1.21–5.45) |

For each of the sub-domains of the JPCA-SF, modified Poisson regression models were fitted with adjustment for age, gender, education, household income, dialysis duration, modified Carlson Index, and the participating facilities.
JPCAT-SF subscale score quartiles:
First contact: Q1, 0.0–37.5; Q2, 50.0–62.5; Q3, 75.0; Q4, 87.5–100.0,
Longitudinality: Q1, 0.0–37.5; Q2, 50.0–62.5; Q3, 75.0; Q4, 87.5–100.0,
Coordination: Q1, 0.0–50.0; Q2, 62.5–75.0; Q3, 87.5; Q4, 100.0,
Comprehensiveness (Services Available): Q1, 0.0–37.5; Q2, 50.0; Q3, 62.5–75.0; Q4, 87.5–100.0,
Comprehensiveness (Services Provided): Q1, 0.0; Q2, 12.5–25.0; Q3, 37.5–50.0; Q4, 62.5–100.0,
Community Orientation: Q1, 0.0–37.5; Q2, 50.0; Q3, 62.5–75.0; Q4, 87.5–100.0.

Supplemental Table 1 shows the continuous relationship between the JPCAT-SF score and ACP participation among patients who reported having a USC. Greater scores on first contact, longitudinality, comprehensiveness (services provided), and community orientation were associated with greater ACP participation.

### Quality of person-centered care by dialysis facility

Among those with a USC, the mean total JPCAT-SF score was 59.8 (SD, 14.4). Figure 3 shows the predicted means of the JPCAT-SF total score by facility, which corresponds to the mean JPCAT-SF total score if all patients in the analysis population received care at a specific facility. Supplementary Table 2 shows the association of the JPCAT-SF total score with the dialysis facility and other covariates. A difference of 12 points was found between the facilities with the highest and lowest total scores.

**Fig 3.**
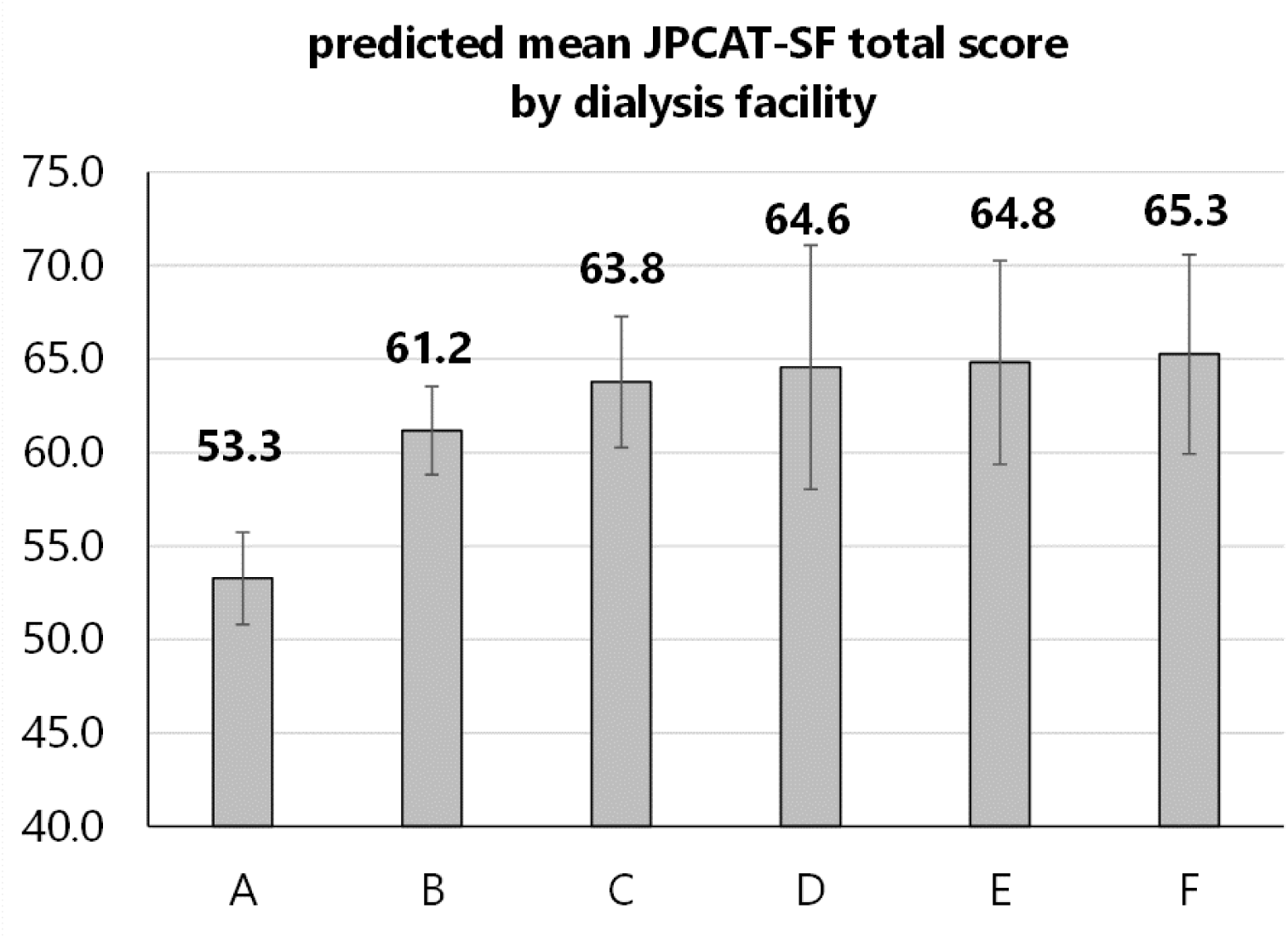
Predicted mean of the JPCAT-SF total scores calculated by participating dialysis facilities (n = 395). Adjusted total scores of the JPCAT-SF scores are predicted using the general linear model displayed in Supplementary Table 3. The left vertical axis indicates the adjusted total score of the JPCAT-SF. Error bars indicate 95% confidence intervals. Gray bars indicate point estimates. For example, if the entire analyzed population belongs to the participating facility A, the predicted mean total score of the JPCAT-SF would be 53.3 (95% confidence interval, 50.8–55.7).

## Discussion

In this study, we showed that a higher level of person-centered care, as measured by the JPCAT-SF, was associated with a higher likelihood of ACP participation among patients on maintenance hemodialysis. This association was consistent across most of the subdomains of the scale (i.e., first contact, longitudinality, comprehensiveness (services provided), and community orientation). Furthermore, the variation in the level of person-centered care across dialysis facilities demonstrated the potential for improving care on a facility-level basis.

The observed relationship between better quality of person-centered care and greater ACP participation in this study confirms the theoretical importance of the person-centered approach in ACP among patients with kidney failure, as described in a previous review and synoptic articles. ^10,12,20^ First, the person-centered approach emphasized in previous studies, such as exploring understanding of patients’ conditions and incorporating their values and preferences about care, were largely grounded from theoretical papers for practicing ACP. ^10,11^ For example, dialysis providers were expected to initiate ACP discussions with patients with kidney failure through care triggered by day-to-day communication, with a focus on building rapport and shared decision-making such as by paying attention to conversations and relationships grounded in daily life. ^20^ However, the studies that examined ACP participation that was associated with day-to-day person-centered care were reported only in the non-kidney primary care outpatient setting and in home medical care settings. ^15,16^ Second, although one study quantitatively evaluated nephrologists’ person-centered care toward patients with chronic kidney disease (CKD), ^13^ the measures associated with it were confined to physician–patient relationships (degree of reliance on nephrologists for care) and were not directed toward decision-making processes such as ACP. Consequently, our study is the first to quantitatively demonstrate the potential for greater levels of day-to-day person-centered care for patients undergoing dialysis to foster favorable physician–patient interactions ^7^ and increase the opportunity for ACP participation.

This study has several implications for dialysis providers regarding person-centered care and ACP participation. First, the frequent proportion of patients (13.5%) who did not consider their dialysis facility as a USC provides an opportunity to investigate whether its reasons are attributed to their dialysis provider or not.

This is underscored by the fact that the rate of ACP participation among respondents who reported not having a USC is the lowest. Patients who responded with not having a USC among American adult general health care users reportedly perceived insufficient person-centered communication between physician and patient. ^29^ Given that many patients on dialysis seek primary care from their nephrologists ^14^ and that greater reliance on nephrologist care is found among patients who perceive they receive such adequate care, ^13^ dialysis providers may need to pay close attention to the patient’s needs. An alternative explanation is that these patients may perceive their dialysis facility’s care as less person-centered because they may already have established relationships with other specialists providing necessary care for them. ^13^ Therefore, their relationship with their dialysis physician may be relatively weaker.

Second, significant relationships between total scores and most subdomains of person-centered care with ACP participation provide opportunities for dialysis providers to focus on specific day-to-day care that can serve as clues for ACP discussions.

First contact, which ensures care when a patient becomes suddenly sick, allows dialysis providers more opportunity to understand patients’ preferences and values on treatment for a critical condition that makes the patient unable to express their wishes, and consequently, to discuss ACP. Thus, first contact supports Kidney Disease: Improving Global Outcomes’s advocacy that ACP should be a part of the process that occurs when patients experience sentinel events such as hospitalization or acute illness. ^30^ Longitudinality, which reflects understanding patients as whole persons (holistic) and capturing their priorities, may facilitate personalized discussions about future health conditions they may experience and related treatment choices through a focus on life-oriented conversations and relationships. ^20^ Indeed, longitudinality is an essential component of person-centeredness in patients with kidney failure; ^31^ a systematic review indicated that lack of it poses a barrier to person-centered care for patients with CKD. In addition to elaborating on interactions with patients, efforts to understand patients and their priorities through active collaboration and communication between dialysis physicians and nurses may also promote ACP participation. ^32^ Comprehensiveness (services provided), which guarantees advice on over-the-counter medications and media health information, may promote ACP participation through a dialogue with their dialysis providers regarding their concerns about their future health condition and negative preconceptions about treatment, which are augmented by lay stories and gathered information from mass media. ^33,34^ Community orientation, which reflects a community-oriented approach, may enable the presentation of pragmatic options for end-of-life care through the effective use of healthcare resources where patients on dialysis live. ^16^

Third, observed differences in the level of person-centered care among dialysis facilities demonstrated in this study suggest an opportunity to consider quality improvement on a facility level. This difference may reflect differences in facilities’ care policies or structural resources. Particularly, the largest difference in total score between facilities may exceed a minimally important difference (12.0 > 0.5 x 14.4 = 7.2, given that the rule of thumb for a minimally important difference is 0.5 x SD of the instrument employed ^35^).

This study has several strengths. First, the multicenter study design with high participation rates ensures the generalizability of our findings. Second, we were able to demonstrate the association between person-centered care and ACP participation while adjusting for case-mix differences. The interesting finding of higher comorbidity scores (i.e., modified CCI) being associated with increased ACP participation raises a reality that dialysis providers prepare for ACP by expecting relatively short prognoses based on accumulated comorbidities.

At the same time, our study has several limitations. First, we did not inquire about detailed life-extending treatments such as discontinuation of dialysis therapy or other invasive treatments. ^2,36^ Indeed, previous studies pointed out that patients with kidney failure who completed advance directives typically do not mention discontinuation of dialysis therapy within them. ^20^ Second, it might be obvious that person-centered care, as measured by the JPCAT-SF, and ACP participation has an association because the item about the availability of counseling on one’s end of life is included in the JPCAT-SF subdomain of comprehensiveness (service available). However, those that were actually associated were other sub-domains. The “end of life in one’s own way,” as measured by the comprehensiveness (service available) subdomain, is not limited to medical care, and perhaps patients on dialysis may be discussing non-ACP aspects of their lives, such as the achievement of their remaining life goals, with their dialysis providers. Third, there are unmeasured confounding factors such as the presence of family members and the duration of the relationship with the healthcare provider. The role of family members is important for person-centered care. However, whether family members facilitate ACP discussions between patients and their dialysis providers remains unclear because the percentage of patients’ families being able to predict their patients’ ACP preferences was no more accurate than would be expected by chance in Japan, ^37^ and the role of their family members in ACP has been noted to vary from patient to patient. ^7^ Additionally, longer relationships with the same dialysis provider may result in receiving better person-centered care. However, although we adjusted for dialysis vintage as a surrogate for the length of the relationship with their dialysis provider, we could not find an association between dialysis vintage and ACP participation.

## Conclusion

The current study demonstrated that higher levels of person-centered care were associated with greater ACP participation among patients undergoing hemodialysis.

Additionally, person-centered care varied by facility. Dialysis providers should strive to treat patients as whole persons and provide as much person-centered day-to-day care as possible through a good provider-patient relationship.

## Supplementary Material

Item S1. Items and responses for the JPCAT-SF

Item S2. Description of the concepts of the JPCAT-SF subdomains

Supplementary Table 1. Association of ACP participation with the JPCAT-SF continuous scores (n = 395)

Supplementary Table 2. Association of the JPCAT-SF total score with facility and covariates (n = 395)

## Article Information

### Authors’ Contributions

Research idea and study design: YK, NK, TO; data acquisition: YK, MU, RI, AK, TT, MM, YM, TS; data analysis/interpretation: TA, YK, NK; statistical analysis: YK, NK; supervision or mentorship: NK, TO. Each author contributed important intellectual content during manuscript drafting or revision, agreed to be personally accountable for the individual’s own contributions, and ensured that questions pertaining to the accuracy or integrity of any portion of the work, even one in which the author was not directly involved, were appropriately investigated and resolved, including documentation in the literature, if appropriate.

### Support

This study was supported by JSPS KAKENHI (grant numbers: JP19KT0021).

## Data Availability

All data produced in the present work are contained in the manuscript.

## Acknowledgments

The authors greatly thank the following researchers, research assistants, and medical staff members for their assistance in collecting the questionnaire-based and clinical information used in this study: Ms. Aki Tairaku (Shin-Yurigaoka General Hospital, Kawasaki-City, Kanagawa); Tetuo Ueki, MD, Akio Munakata, MD, Yoshihiko Watanabe, MD (Munakata Clinic, Mobara-City, Chiba); Ms. Yayoi Takanashi, Reiji Masaki, NP, Tomohiko Inoue, MD, Shinnosuke Sugihara, MD, Kanako Nagaoka, MD and Hiroshi Kuji, MD (Kameda Medical Center, Kamogawa-City, Chiba); Kenji Yamaguchi, MD (Awa Regional Medical Center, Tateyama-City, Chiba); Ms. Miyuki Sato (Fukushima Medical University Hospital, Fukushima-City, Fukushima).

## References

1. Brinkman-Stoppelenburg A, Rietjens JAC, van der Heide A. The effects of advance care planning on end-of-life care: a systematic review. Palliat Med. 2014;28(8):1000–1025. doi:10.1177/0269216314526272

2. Rietjens JAC, Sudore RL, Connolly M, et al. Definition and recommendations for advance care planning: an international consensus supported by the European Association for Palliative Care. Lancet Oncol. 2017;18(9):e543–e551. doi:10.1016/S1470-2045(17)30582-X

3. Molloy DW, Guyatt GH, Russo R, et al. Systematic implementation of an advance directive program in nursing homes: a randomized controlled trial. JAMA. 2000;283(11):1437–1444. doi:10.1001/jama.283.11.1437

4. Wright AA, Zhang B, Ray A, et al. Associations between end-of-life discussions, patient mental health, medical care near death, and caregiver bereavement adjustment. JAMA. 2008;300(14):1665–1673. doi:10.1001/jama.300.14.1665

5. O’Malley AJ, Caudry DJ, Grabowski DC. Predictors of nursing home residents’ time to hospitalization. Health Serv Res. 2011;46(1 Pt 1):82–104. doi:10.1111/j.1475-6773.2010.01170.x

6. USRDS. End Stage Renal Disease: Chapter 6 Mortality. 2022 Annual Report. Published 2022. https://usrds-adr.niddk.nih.gov/2022/end-stage-renal-disease/6-mortality

7. Davison SN. Facilitating advance care planning for patients with end-stage renal disease: the patient perspective. Clin J Am Soc Nephrol. 2006;1(5):1023–1028. doi:10.2215/CJN.01050306

8. Sellars M, Clayton JM, Morton RL, et al. An Interview study of patient and caregiver perspectives on advance care planning in ESRD. Am J Kidney Dis. 2018;71(2):216–224. doi:10.1053/j.ajkd.2017.07.021

9. Goff SL, Unruh ML, Klingensmith J, et al. Advance care planning with patients on hemodialysis: an implementation study. BMC Palliat Care. 2019;18(1):64. doi:10.1186/s12904-019-0437-2

10. O’Halloran P, Noble H, Norwood K, et al. Advance Care planning with patients who have end-stage kidney disease: a systematic realist review. J Pain Symptom Manage. 2018;56(5):795–807.e18. doi:10.1016/j.jpainsymman.2018.07.008

11. O’Hare AM. Patient-Centered care in renal medicine: five strategies to meet the challenge. Am J Kidney Dis. 2018;71(5):732–736. doi:10.1053/j.ajkd.2017.11.022

12. Morton RL, Sellars M. From Patient-centered to person-centered care for kidney diseases. Clin J Am Soc Nephrol. 2019;14(4):623–625. doi:10.2215/CJN.10380818

13. Barrett TM, Green JA, Greer RC, et al. Advanced CKD Care and decision making: which health care professionals do patients rely on for CKD Treatment and advice? Kidney Med. 2020;2(5):532–542.e1. doi:10.1016/j.xkme.2020.05.008

14. Wang V, Diamantidis CJ, Wylie J, Greer RC. Minding the gap and overlap: a literature review of fragmentation of primary care for chronic dialysis patients. BMC Nephrol. 2017;18(1):274. doi:10.1186/s12882-017-0689-0

15. Aoki T, Miyashita J, Yamamoto Y, et al. Patient experience of primary care and advance care planning: a multicentre cross-sectional study in Japan. Fam Pract. 2017;34(2):206–212. doi:10.1093/fampra/cmw126

16. Hayashi S, Shirahige Y, Fujioka S, et al. Relationship between patient-centred care and advance care planning among home medical care patients in Japan: the Zaitaku evaluative initiatives and outcome study. Fam Pract. 2023;40(2):211–217. doi:10.1093/fampra/cmac062

17. Ladin K, Neckermann I, D’Arcangelo N, et al. Advance Care planning in older adults with CKD: patient, care partner, and clinician perspectives. J Am Soc Nephrol. 2021;32(6):1527–1535. doi:10.1681/ASN.2020091298

18. O’Hare AM, Kurella Tamura M, Lavallee DC, et al. Assessment of Self-reported prognostic expectations of people undergoing dialysis: United States renal data system study of treatment preferences (USTATE). JAMA Intern Med. 2019;179(10):1325–1333. doi:10.1001/jamainternmed.2019.2879

19. Houben CHM, Spruit MA, Groenen MTJ, Wouters EFM, Janssen DJA. Efficacy of advance care planning: a systematic review and meta-analysis. J Am Med Dir Assoc. 2014;15(7):477–489. doi:10.1016/j.jamda.2014.01.008

20. Davison SN, Torgunrud C. The creation of an advance care planning process for patients with ESRD. Am J Kidney Dis. 2007;49(1):27–36. doi:10.1053/j.ajkd.2006.09.016

21. Aoki T, Fukuhara S, Yamamoto Y. Development and validation of a concise scale for assessing patient experience of primary care for adults in Japan. Fam Pract. 2020;37(1):137–142. doi:10.1093/fampra/cmz038

22. Aoki T, Inoue M, Nakayama T. Development and validation of the Japanese version of Primary Care Assessment Tool. Fam Pract. 2016;33(1):112–117. doi:10.1093/fampra/cmv087

23. Shi L, Starfield B, Xu J. Validating the Adult Primary Care Assessment Tool. J Fam Pract. 2001;50(2).

24. Hudon C, Fortin M, Haggerty JL, Lambert M, Poitras ME. Measuring patients’ perceptions of patient-centered care: a systematic review of tools for family medicine. Ann Fam Med. 2011;9(2):155–164. doi:10.1370/afm.1226

25. Hemmelgarn BR, Manns BJ, Quan H, Ghali WA. Adapting the Charlson Comorbidity Index for use in patients with ESRD. Am J Kidney Dis. 2003;42(1):125–132. doi:10.1016/s0272-6386(03)00415-3

26. Zou G. A modified poisson regression approach to prospective studies with binary data. Am J Epidemiol. 2004;159(7):702–706. doi:10.1093/aje/kwh090

27. Aoki T, Sugiyama Y, Mutai R, Matsushima M. Impact of Primary care attributes on hospitalization during the COVID-19 Pandemic: a nationwide prospective cohort study in Japan. Ann Fam Med. 2023;21(1):27–32. doi:10.1370/afm.2894

28. Williams R. Using the margins command to estimate and interpret adjusted predictions and marginal effects. Stata J. 2012;12(2):308–331. doi:10.1177/1536867X1201200209

29. Finney Rutten LJ, Agunwamba AA, Beckjord E, Hesse BW, Moser RP, Arora NK. The Relation between having a usual source of care and ratings of care quality: does patient-centered communication play a role? J Health Commun. 2015;20(7):759–765. doi:10.1080/10810730.2015.1018592

30. Davison SN, Levin A, Moss AH, et al. Executive summary of the KDIGO Controversies Conference on Supportive Care in Chronic Kidney Disease: developing a roadmap to improving quality care. Kidney Int. 2015;88(3):447–459. doi:10.1038/ki.2015.110

31. de Jong Y, van der Willik EM, Milders J, et al. Person centred care provision and care planning in chronic kidney disease: which outcomes matter? A systematic review and thematic synthesis of qualitative studies: care planning in CKD: which outcomes matter? BMC Nephrol. 2021;22(1):309. doi:10.1186/s12882-021-02489-6

32. O’Hare AM, Szarka J, McFarland LV, et al. Provider Perspectives on advance care planning for patients with kidney disease: whose job is it anyway? Clin J Am Soc Nephrol. 2016;11(5):855–866. doi:10.2215/CJN.11351015

33. Seah AST, Tan F, Srinivas S, Wu HY, Griva K. Opting out of dialysis – Exploring patients’ decisions to forego dialysis in favour of conservative non-dialytic management for end-stage renal disease. Health Expect. 2015;18(5):1018–1029. doi:10.1111/hex.12075

34. Lin CC, Chen MC, Hsieh HF, Chang SC. Illness representations and coping processes of Taiwanese patients with early-stage chronic kidney disease. J Nurs Res. 2013;21(2):120–128. doi:10.1097/jnr.0b013e3182921fb8

35. Norman GR, Sloan JA, Wyrwich KW. Interpretation of changes in health-related quality of life: the remarkable universality of half a standard deviation. Med Care. 2003;41(5):582–592. doi:10.1097/01.MLR.0000062554.74615.4C

36. Wong SPY, Prince DK, Kurella Tamura M, et al. Value Placed on comfort vs life prolongation among patients treated with maintenance dialysis. JAMA Intern Med. 2023;183(5):462–469. doi:10.1001/jamainternmed.2023.0265

37. Miura Y, Asai A, Matsushima M, et al. Families’ and physicians’ predictions of dialysis patients’ preferences regarding life-sustaining treatments in Japan. Am J Kidney Dis. 2006;47(1):122–130. doi:10.1053/j.ajkd.2005.09.030

