## Supplementary Files for "Association Between Person-Centered Care Quality and Advance Care Planning Participation in Patients Undergoing Hemodialysis: A Multicenter Cross-Sectional Study"

**Supplementary materials**

### **Supplementary Item S1. Items and responses for the JPCAT-SF [1]**

Questionnaires in Japanese version are available from the following website (<https://bfffe681-45f7-48c3-aa93-2f67612434a5.filesusr.com/ugd/6c0e9c_7c80f3e3ce6e45eebb5a3aedfb9a2800.pdf>).

| **Instruction sentences** | **(English: “Check the box that best fits each question.”)** |
| --- | --- |
| Question 1 | (English: “When your Primary Care Practice is closed on Saturday and Sunday and you get sick, would someone from there see you the same day?”) |
| Response to Question 1 | (English: “Strongly agree/Somewhat agree/Not sure/Somewhat disagree/Strongly disagree ”) |
| Question 2 | (English: “When your Primary Care Practice is closed and you get sick during the night, would someone from there see you that night?”) |
| Response to Question 2 | (English: “Strongly agree/Somewhat agree/Not sure/Somewhat disagree/Strongly disagree ”) |
| Question 3 | (English: “Does your Primary Care Physician (PCP) know you very well as a person, rather than as someone with a medical problem?”) |
| Response to Question 3 | (English: “Strongly agree/Somewhat agree/Not sure/Somewhat disagree/Strongly disagree ”) |
| Question 4 | (English: “Does your PCP know what problems are most important to you?”) |
| Response to Question 4 | (English: “Strongly agree/Somewhat agree/Not sure/Somewhat disagree/Strongly disagree ”) |
| Question 5 | (English: “Have you ever had a visit to a specialist or special service of any kind?”) |
| Response to Question 5 | (English: “Yes/No or Not sure”) |
| Question 6 | (English: “Did your PCP suggest you go to the specialist or special service?”) |
| Response to Question 6 | (English: “Strongly agree/Somewhat agree/Not sure/Somewhat disagree/Strongly disagree ”) |
| Question 7 | (English: “Did your PCP discuss with you the different places you could have visited to get help with that problem?”) |
| Response to Question 7 | (English: “Strongly agree/Somewhat agree/Not sure/Somewhat disagree/Strongly disagree ”) |
| Question 8 | (English: “Please indicate whether it is available at your PCP’s office. Counselling related to abuse”) |
| Response to Question 8 | (English: “Strongly agree/Somewhat agree/Not sure/Somewhat disagree/Strongly disagree ”) |
| Question 9 | (English: “Please indicate whether it is available at your PCP’s office. Counselling related to personal preferences about end-of-life issues”) |
| Response to Question 9 | (English: “Strongly agree/Somewhat agree/Not sure/Somewhat disagree/Strongly disagree ”) |
| Question 10 | (English: “In visits to your PCP, are any of the following subjects discussed with you? Advice about over-the-counter medications.”) |
| Response to Question 10 | (English: “Strongly agree/Somewhat agree/Not sure/Somewhat disagree/Strongly disagree ”) |
| Question 11 | (English: “In visits to your PCP, are any of the following subjects discussed with you? Advice about medical information in the media: on TV, in the newspaper, etc.”) |
| Response to Question 11 | (English: “Strongly agree/Somewhat agree/Not sure/Somewhat disagree/Strongly disagree ”) |
| Question 12 | (English: “Does your PCP investigate whether the available health care is meeting the needs of the community?”) |
| Response to Question 12 | (English: “Strongly agree/Somewhat agree/Not sure/Somewhat disagree/Strongly disagree ”) |
| Question 13 | (English: “Does your PCP investigate the concerns people have about health problems in your community?”) |
| Response to Question 13 | (English: “Strongly agree/Somewhat agree/Not sure/Somewhat disagree/Strongly disagree ”) |

Each domain consists of the following items:

First contact domain - Questions 1 and 2

Longitudinality domain - Questions 3 and 4

Coordination domain - Questions 5, 6, and 7

Comprehensiveness (services available) domain - Questions 8 and 9

Comprehensiveness (services provided) domain - Questions 10 and 11

Community orientation domain - Questions 12 and 13

#### Scoring

For each item, participants were asked to respond on a 5-point Likert scale ranging from *strongly disagree* to *strongly agree*. Each response was converted to an item score ranging from 0 to 4. The domain scores were calculated by multiplying the average of the item scores in the same domain by 25 (i.e., ranging from 0 to 100), with higher scores indicating better performance. In the coordination domain, which asks about experiences with referrals to a specialist, respondents who had never seen a specialist were given 50 points (the midpoint of all possible scores). The total score was the average of the six domain scores and represented an overall measure of the patient experience of primary care.

**Reference**

1.	Aoki T, Fukuhara S, Yamamoto Y. Development and validation of a concise scale for assessing patient experience of primary care for adults in Japan. *Fam Pract.* 2020;37(1):137-142. doi: 10.1093/fampra/cmz038

**Supplementary Item S2. Description of the concepts of the JPCAT-SF subdomains**

The JPCAT-SF is a short version of the original 29-item JPCAT,^1^ which is an adaptation to the Japanese culture of the Primary Care Assessment Tool (PCAT) that was designed to measure the experience of adult patients in primary care.^2^ In an outpatient setting, the JPCAT-SF has been shown to have good internal consistency reliability (Cronbach’s α = 0.77 for the total score, Cronbach’s α > 0.76 for each domain score) and excellent criterion validity (Pearson correlation coefficient with the original 29-item JPCAT and the overall rating for usual care facilities: 0.94 and 0.43, respectively).^3^

First contact

Care is first sought from a primary care provider when a new health or medical need arises. The service should also be accessible and usable by the population as a new need or problem arises.^4^ First contact is closely related to “access to care,” a domain of person-centered care characterized by the timely availability of care that is tailored to the patient.^5^ The JPCAT-SF mainly measures patient experience related to off-hours care in primary care.^4^

Longitudinality

It refers to the longitudinal use of usual sources of care regardless of illness or injury.^4^ Longitudinality is supported by one of the principles for person-centeredness, namely the consideration of the “patient as a unique person,” i.e., the primary care physician’s recognition of the patient’s uniqueness (individual needs, preferences, values, beliefs, concerns, etc.).^5^ The JPCAT-SF mainly measures whether a patient feels that their primary care physician recognizes them as a whole person.^4^

Coordination

The essence of coordination is the availability of information about past and existing problems and services and the recognition of that information in relation to a current care need.^4^ It relates to “coordination and continuity of care,” which is an enabler of person-centered care, i.e., facilitation of care that is well-coordinated and continuous.^5^ The JPCAT-SF mainly measures patient experience regarding past referrals to a specialist.^4^

Comprehensiveness (services available)

It refers to the availability of a wide range of services by a primary care provider and their appropriateness for a spectrum of needs for all but the most uncommon problems.^4^

Under “services available,” the JPCAT-SF mainly measures whether a patient feels they can receive care for mental health, dementia, and advanced care planning, if necessary.^4^

Comprehensiveness (services provided)

It includes appropriate advice on daily lifestyle habits, self-medication, and health literacy.^4,6^ It is underpinned by patient empowerment, an activity of person-centeredness, in which a primary care physician recognizes and actively supports a patient's ability and responsibility to self-manage their illness.^5^ The JPCAT-SF mainly measures patient experience in terms of whether they received such appropriate advice.

Community orientation

It refers to care that is delivered in the context of the community^4^ and is considered as a derivative domain of principles of primary care.^1^

The JPCAT-SF mainly measures patient experience regarding home visits and whether a patient feels that their primary care physician is interested not only in their individual health problem but also in problems in the community.^4^

**References**

1.	Aoki T, Inoue M, Nakayama T. Development and validation of the Japanese version of Primary Care Assessment Tool. *Fam Pract.* 2016;33(1):112-117. doi: 10.1093/fampra/cmv087

2.	Shi L, Starfield B, Xu J. Validating the adult primary care assessment tool. *J Fam Pract.* 2001;50(2):161-161. doi:

3.	Aoki T, Fukuhara S, Yamamoto Y. Development and validation of a concise scale for assessing patient experience of primary care for adults in Japan. *Fam Pract.* 2020;37(1):137-142. doi: 10.1093/fampra/cmz038

4.	Kaneko M, Aoki T, Goto R, Ozone S, Haruta J. Better Patient Experience is Associated with Better Vaccine Uptake in Older Adults: Multicentered Cross-sectional Study. *J Gen Intern Med.* 2020;35(12):3485-3491. doi: 10.1007/s11606-020-06187-1

5.	Scholl I, Zill JM, Härter M, Dirmaier J. An integrative model of patient-centeredness - a systematic review and concept analysis. *PLoS One.* 2014;9(9):e107828-e107828. doi: 10.1371/journal.pone.0107828

6.	Aoki T, Yamamoto Y, Fukuhara S. Comparison of Primary Care Experience in Hospital-Based Practices and Community-Based Office Practices in Japan. *Ann Fam Med.* 2020;18(1):24-29. doi: 10.1370/afm.2463

### **Supplementary Table 1. Association of ACP participation with the JPCAT-SF continuous scores (n = 395)**

|  | Adj. PR, point estimate (95% CI) |  |  |  |  |  |  |
| --- | --- | --- | --- | --- | --- | --- | --- |
|  | Total Score | First Contact | Longitudinality | Coordination | Comprehensiveness (Services Available) | Comprehensiveness (Services Provided) | Community Orientation |
| JPCAT-SF scores |  |  |  |  |  |  |  |
| per 10-pt increase | 1.19 (1.07–1.33) | 1.09 (1.006–1.17) | 1.09 (1.002–1.18) | 1.06 (0.991–1.13) | 1.03 (0.95–1.1) | 1.06 (1.01–1.11) | 1.07 (1.002–1.15) |

For each of the sub-domains of the JPCA-SF, modified Poisson regression models were fitted with adjustment for age, gender, education, household income, dialysis duration, modified Carlson Index, and the participating facilities.

### **Supplementary Table 2. Association of the JPCAT-SF total score with facility and covariates (n = 395)**

|  | Adj. mean difference, point estimate (95% CI) | P-value |
| --- | --- | --- |
| Facility |  |  |
| *facility A* | Reference |  |
| *facility B* | 7.9 (4.44–11.37) | <0.001 |
| *facility C* | 10.5 (6.23–14.76) | <0.001 |
| *facility D* | 11.28 (4.3–18.27) | 0.002 |
| *facility E* | 11.54 (5.57–17.51) | <0.001 |
| *facility F* | 11.98 (6.11–17.86) | <0.001 |
| Age, per 10-yr increase | 0.27 (-0.91–1.44) | 0.652 |
| Women vs. Men | 1.44 (-1.53–4.41) | 0.342 |
| Education |  |  |
| *Junior high school graduate or less* | Reference |  |
| *High school graduate* | −3.6 (−7.17–−0.04) | 0.048 |
| *University/Graduate school graduate* | −5.54 (−10.65–−0.43) | 0.034 |
| *Others* | −2.23 (−7.1–2.63) | 0.367 |
| Household Income |  |  |
| <3,000,000 yen | Reference |  |
| 3,000,000 to <5,000,000 yen | −0.21 (−3.58–3.15) | 0.901 |
| ≥5,000,000 yen | −1.82 (−5.59–1.96) | 0.344 |
| Dialysis vintage, per 1-yr increase | −0.11 (−0.28–0.07) | 0.249 |
| Modified Charlson score, per 1-pt increase | 0.55 (−0.19–1.29) | 0.142 |

A general linear model was fit with adjustment for age, gender, education, household income, dialysis duration, modified Carlson Index, and the participating facilities.
